# Interpreting Health Differences between Self-reported Black and White Children in U.S.: Insights from a Methodological Perspective

**DOI:** 10.1101/2024.10.01.24314712

**Authors:** Fan Nils Yang, Jeff H. Duyn, Weizhen Xie

## Abstract

Understanding health differences among racial groups in child development is crucial for addressing inequalities that may affect various aspects of a child’s life. However, factors such as household and neighborhood socioeconomic status (SES) often covary with health differences between races, making it challenging to accurately reveal these differences using conventional covariate-control methods such as multiple regression. Alternative methods, such as Propensity Score Matching (PSM), may provide better covariate control. Supporting this notion, we found that PSM is more sensitive than regression-based methods in detecting health differences between self-reported Black and White children across a wide range of behavioral and neural measurements in the ABCD (5636 White, 1350 Black). Puberty status, an index of physical maturation, emerged as the largest difference between races and mediated the health differences between races on the majority of behavioral and neural variables. These findings highlight the importance of controlling for pubertal status and using more effective covariate-control methods to accurately represent health differences between Black and White children.

## Introduction

Nearly half of American adolescents in 2019 identified as belonging to a racial or ethnic minority (Bureau, 2020), reflecting the increasing diversity of the U.S. population. Understanding health differences between races and ethnicities in adolescents is essential for designing and delivering services that are accessible, equitable, and culturally attuned to this population (Karcher et al., 2022). However, pinpointing health differences between races in adolescents presents a challenge, considering numerous covariates of race and ethnicity (e.g., SES and geographic variations) that can confound or mediate these differences. For instance, while Black children have been found to sleep less compared to White children (Giddens et al., 2022), controlling for additional SES variables beyond household income (e.g., parents’ education level) may reduce this difference (Yang et al., 2022). Furthermore, the variability in the selected covariates and the use of potentially suboptimal covariate-control methods in some prior research may further add to this uncertainty (Dick et al., 2021), especially when accessing neurobiological correlates (Dumornay et al., 2023). Nevertheless, there is a critical need to accurately represent health differences among racial groups, as doing so is key to achieving health equity within the diverse U.S. adolescent population.

The longitudinal behavioral and neural data in 11,878 9-10-year-olds from the ABCD study (Casey et al., 2018) present a unique opportunity to systematically address this issue. To this end, our primary objective is to investigate health differences between two major self-report racial groups among the participants in the ABCD study, namely Black and White children. We compared their differences in behavioral and neural measures based on a unified selection of basic covariates across different covariate-control methods (i.e., multiple regression and PSM (Rosenbaum & Rubin, 1983)). These covariates include age, sex, the interaction between age and sex, household income, parents’ education level, Area Deprivation Index (ADI), and study sites. As analysis revealed improved sensitivity of PSM in detecting health differences between races during early adolescence, we noticed that pubertal status exhibited the largest difference (Cohen’s d > 0.60) at baseline.

This finding further prompted us to investigate puberty status as an essential mediator of health differences between races, encompassing aspects of physical and mental health, as well as brain health. We found that many health-related differences between self-reported Black and White children are mediated by their differing rates of pubertal development. Adjusting for between-group differences in pubertal status reduced the health differences observed between Black and White children. By clarifying this relationship, we can better interpret racial differences in adolescents and promote educational programs that are tailored to children at different stages of pubertal development to help address these differences.

## Materials and Methods

### Data source

Data used in the current study were from the ABCD data release 5.1 (2023) (https://abcdstudy.org), which includes behavioral and neural data from 11,878 9-10-year-olds collected at baseline, 1-year (FL1), 2-year (FL2), 3-year (FL3) and 4-year follow-ups (FL4). Detailed protocols and designs have been described previously on the website. Informed consent from the primary caregiver and assent from the children were obtained before the study. This project was approved by institutional review boards (IRB) at the University of California, San Diego as well as at each local site (21 in total). Participants were recruited using stratified sampling to reflect the diversity of the U.S. population. Participants who were excluded from data analysis had missing data for the seven basic covariates (n = 971, out of 7,957 Black or White children). To study health differences between Black and White children, 6,986 (5,636 White, 1,350 Black) out of the entire 11,878 participants were included in this study.

### Behavioral measurements

The independent variable is Children’s race (Black and White), obtained by the following question answered by their parents/caregivers, “Race Ethnicity (Child): 1 = White; 2 = Black; 3 = Hispanic; 4 = Asian; 5 = Other”. We chose Black and White children because these two racial groups comprise the majority (∼80%) of the population in the ABCD study.

Based on prior research, seven basic covariates are included to control for individual differences due to age, sex, socioeconomic status, and study sites (geographic differences) (Yang et al., 2022). They capture a child’s age as months, sex at birth, the interaction between age and sex, study sites (21 sites), family-level socioeconomic status: family income (10 levels, from under $5000 per year to more than $200,000 per year) and highest educational level of caregiver (22 levels, from never attended to doctoral degree), and environment-level socioeconomic status: area deprivation index (ADI, 1-100, lower score means less deprivation). These covariates were selected because they encompass key demographic and socioeconomic factors that could potentially influence health-related outcomes, such as brain morphology and physical development (Rakesh et al., 2022). Ultimately, 64 dependent variables or candidate covariates were included for analysis (see https://data-dict.abcdstudy.org/ for data dictionary), with some of them only available at certain data collection points. For example, the NIH toolbox was only measured at baseline, FL2, and FL4. These 64 variables are:

**(i)** 17 Culture & Environment variables: Cognition (wps_ss_sum), Discrimination Measure (dim_y_ss_mean), Neighborhood Safety & Crime (neighborhood_crime_y), Acculturation Heritage (via_ss_hc), Acculturation Mainstream (via_ss_amer), Family Conflict (fes_y_ss_fc_pr), Children’s Report of Parental Behavior (crpbi_y_ss_parent and crpbi_y_ss_caregiver), Neglect (mnbs_ss_mean_all), Parental Monitoring (pmq_y_ss_mean), Peer Network Health (pnh_ss_protective_scale), Peer Influence (peerinfluence_ss_mean), School Grade (sag_grades_last_yr), School Environment (srpf_y_ss_ses), School Involvement (srpf_y_ss_iiss), School Disengagement (srpf_y_ss_dfs), and Prosocial Behavior (psb_y_ss_mean);
**(ii)** 19 Mental Health variables: Brief Problem Monitor Total (bpm_y_scr_totalprob_t), Emotion Regulation Reappraisal (erq_ss_reappraisal_pr), Emotion Regulation Suppression (erq_ss_suppress_pr), NIH Toolbox Positive Affect (poa_y_ss_sum), Mood Mania (sup_y_ss_sum), Prodromal Psychosis Scale (pps_ss_mean_severity), Stress Life Events (ple_y_ss_total_number), Behavioral Inhibition (bis_y_ss_bis_sum), Behavioral Approach (bis_y_ss_bas_rr, bis_y_ss_bas_drive, and bis_y_ss_bas_fs), Impulsivity Behaviour (upps_y_ss_negative_urgency, upps_y_ss_positive_urgency, upps_y_ss_lack_of_perseverance, upps_y_ss_sensation_seeking, and upps_y_ss_lack_of_planning) and behavior problems (cbcl_scr_syn_internal_t, cbcl_scr_syn_external_t, and cbcl_scr_syn_totprob_t);
**(iii)** 12 Neurocognition variables: Cash Choice Task (cash_choice_task), Flanker task (flkr_scr_medrt_congruent and flkr_scr_medrt_incongruent), Game of Dice (gdt_scr_expressions_net_score), Little Man Task (lmt_scr_efficiency), NIH Toolbox Crystalized Intelligence (nihtbx_cryst_fc), NIH Toolbox Fluid Intelligence (nihtbx_fluidcomp_fc), NIH Toolbox Total Intelligence (nihtbx_totalcomp_fc), NIH Toolbox Verbal Learning Short Delay (pea_ravlt_sd_trial_i_tc), NIH Toolbox Verbal Learning Long Delay (pea_ravlt_ld_trial_vii_tc), Mental Arithmetic (smarte_ss_all_total_corr), and WISC-V Matrix Reasoning (pea_wiscv_tss).
**(iv)** two screen usage variables (stq_y_ss_weekday) and (stq_y_ss_weekend).
**(v)** three Biospecimens variables: Sex Hormone DHEA (hormone_scr_dhea_mean), Sex Hormone Estradiol (hormone_scr_hse_mean), and Sex Hormone Testosterone (hormone_scr_ert_mean).
**(vi)** 11 Physical Health variables: Pain Scale (painscale), Activity Involvement Read (sai_read_hrs_wk_y), Activity Involvement Music (sai_lmusic_hrs_day_y), Activity Level (physical_activity1_y and physical_activity2_y), Sleep Disturbance (sds_p_ss_total), Sleep Duration (mctq_sdweek_calc), Chronotype (mctq_msfsc_calc), Waist Size (anthro_waist_cm), BMI, and Pubertal Status (pds_p_ss_female_category_2/pds_p_ss_male_category_2), which is based on different levels of physical maturation, was measured by parent-reported Youth Pubertal Development Scale and Menstrual Cycle Survey. In this survey, individuals rate their child’s development on a four-point Likert scale from “had not begun” to “already complete” with respect to specific physical characteristics (e.g., skin changes, breast development; a subset of the items was administered based on sex). Pubertal Status is a sum score of all the physical characteristics, and it has four levels: 1-pre-puberty, 2-early-puberty, 3-mid-puberty, and 4-post-puberty.

### Neural measurements

All children underwent standardized resting-state fMRI and structural MRI imaging scans at baseline, FL2, and FL4. Acquired images were processed and quality controlled at the Data Analysis, Informatics and Resource Center of the ABCD study (Hagler et al., 2019). Resting-state functional connectivity (rs-FC) of cortical networks was calculated as the average Fisher-transformed correlation between the time courses of each pair of regions within or between 12 cortical networks defined by the Gordon atlas. In addition, rs-FC between the 12 cortical networks and 19 subcortical regions were also calculated. Gray matter volumes (GMV) from 148 regions were extracted based on the Destrieux Parcellation. Subcortical volumes of 30 regions were also calculated based on protocols in the ABCD study. In total, 416 rs-FC and 178 GMV measurements were included.

### Data Analysis

We employed two different methods for controlling covariates. In the regression-based approach, we first regressed out seven basic covariates from the dependent variables in the full sample of 6,986 children. We then compared the standardized mean difference between Black and White children on the resulting residual scores using an independent-sample t-test (Kolisnyk et al., 2023). This method captures the unique variance attributed to race groups and estimates the corresponding effect size in terms of Cohen’s d (Black children - White children). This is analogous to the traditional method of estimating the regression coefficient for a key predictor after accounting for the variance explained by the covariates (Cohen et al., 2002). For the PSM method, we used the ‘MatchIt’ package in R. Children were matched based on the probability of being in a comparison group conditioned on seven basic covariates using logistic regression. Specifically, White children were matched with Black children using one-to-one matching without replacement within a predefined propensity score radius (i.e., caliper = 0.1). Quality-check showed all seven basic covariates were balanced between groups after matching (standardized mean difference lower than 0.05, see **Figure S1**). In the end, 924 pairs were identified. PSM-based Cohen’s d was calculated using an independent sample t-test on these matched pairs. For rs-FC analysis, mean motion and the number of time points remained after preprocessing were added as additional covariates. For GMV analysis, intracranial volumes were added as an additional covariate.

### Mediation analysis

The mediation toolbox (https://github.com/canlab/MediationToolbox) (Wager et al., 2008, 2009) was used to perform all the mediation analyses. Here, in **Figure 2A** the independent variable (X) was the racial group, the dependent variable (Y) was behavioral measurements or brain measurements, and the mediator (M) was pubertal status. The test of mediation involves two linear equations (see equations 1 and 2 below). The path coefficient a reflects the effects of X on M. The path coefficient b effect reflects the effect of M on Y. The coefficient c’ is the direct effect of X on Y after controlling for M. The product a*b (mediation effect/indirect effect) reflects how the association between X and Y changed according to M. d1 and d2 are intercept terms (content). The total effect c (not shown in the equation, c = a*b + c’) is the effect of X on Y without controlling for M. All seven basic covariates were controlled in the mediation analyses. The effect size of meditation was calculated as the beta value of path a*b divided by the beta value of total effect c. The significance of the mediation analyses was estimated using bootstrap sampling with 10,000 random-generated samples on the product of the a and b path coefficients (a*b).

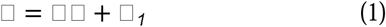

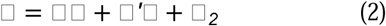

## Results

To study health differences between self-reported Black and White children in the ABCD data release 5.1, seven basic covariates, including age, sex, interaction between age and sex, household income, parents’ education level, ADI, and study sites, were controlled either using the multiple regression method or PSM. After PSM, 924 pairs of Black and White children were identified, and then independent t-tests were performed on these matched pairs (PSM sample n = 1,846). For multiple regression, the basic covariates were first regressed out from the dependent variables, and the residuals were then compared using independent t-tests (regression sample n = 6,986, including 5,636 White, 1,350 Black). See **Table 1 for the demographic and socioeconomic variables of these 6,986 children.**

**Table 1.** Demographic and socioeconomic information for the participants included in the study.

| Characteristic | White | Black | Cohen's D |
| --- | --- | --- | --- |
| Age (month) | 119.11 (7.52) | 118.97 (7.17) | 0.02 |
| Sex | 1.47 (0.50) | 1.51 (0.50) | -0.07 |
| Household Income | 8.17 (1.68) | 5.10 (2.62) | 1.62 |
| Parent Education | 17.11 (3.54) | 14.17 (5.02) | 0.76 |
| ADI | 31.22 (21.98) | 60.97 (32.61) | -1.22 |
| Study Sites | 12.65 (5.76) | 13.02 (5.80) | 0.06 |
| BMI | 17.87 (3.37) | 20.65 (5.30) | -0.73 |
| Pubertal Status | 1.58 (0.78) | 2.28 (0.91) | -0.86 |
| Number of Participants | 5636 | 1350 |  |

Note: ADI: area deprivation index; BMI: body mass index.

### Physical and Mental Health differences between Black and White children at baseline

We first examined health differences in variables depicting a child’s physical and mental health (PSM v.s. Multiple regression). This includes 64 dependent variables from the ABCD dataset, capturing the family/community environment (e.g., parent neglect), mental health (e.g., behavior problems), physical health (e.g., puberty status), technology use (e.g., screen time), neurocognition (e.g., total intelligence), and biospecimen variables (e.g., sex hormones). Among the 41 variables available at baseline, 27 showed significant racial differences based on PSM between self-reported Black and White children, while 26 showed similar significant racial differences based on multiple regression analyses (FDR-corrected p-value < 0.05). For 18 variables with a Cohen’s d value greater than 0.15, the PSM-based effect size is significantly larger than the regression-based effect size, extending outside the 95% confidence interval of the regression-based effect size estimates (**Figure 1A**). This indicates that PSM is more sensitive than the regression-based method in detecting health differences between races. Notably, pubertal status, as an index based on physical maturation measured by the parent-reported Youth Pubertal Development Scale and Menstrual Cycle Survey, emerged as the largest difference between Black and White children (Cohen’s d = 0.64 based on PSM and 0.53 based on regression analysis; **Figure 1A**).

**Figure 1.**
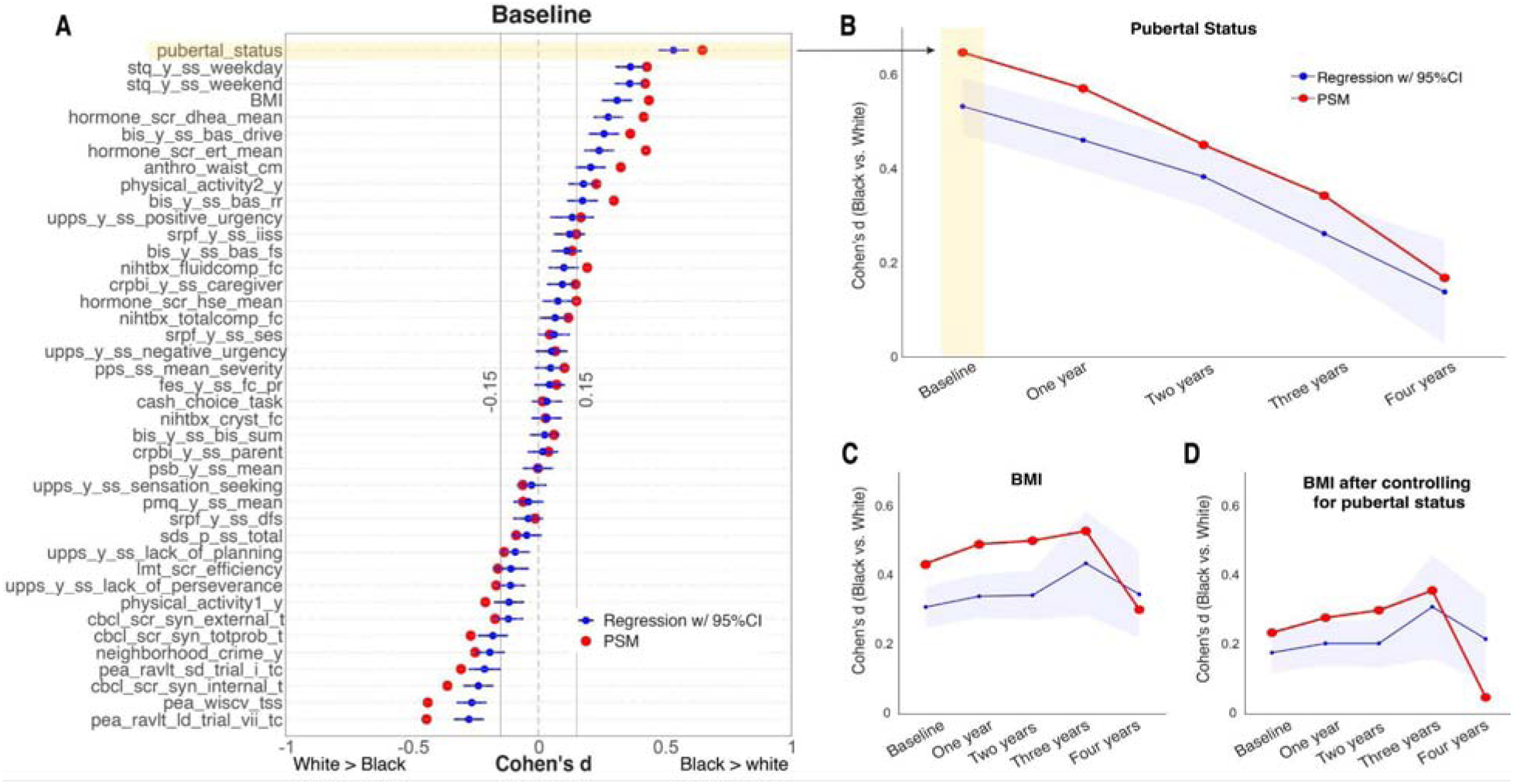
Effect size (Cohen’s d) comparisons between PSM-based (red) and regression-based (blue) methods. (**A**) Cohen’s d values of 41 variables at baseline. For details about variable names, see **Methods**. (**B**) The change of Cohen’s d value of pubertal status across five timepoints. The change of Cohen’s d value of BMI across five timepoints without (**C**) or with (**D**) controlling for pubertal status at each time point.

### Difference of Pubertal status between races and its impact on child development

The gap in pubertal status between Black and White children aligns with past findings (Argabright et al., 2022). We further examined how differences in pubertal status between races change throughout development. Across both the PSM and conventional regression-based methods, the difference in pubertal status between races diminished as children progressed from ages 9-11 to 13-15 over a 4-year longitudinal follow-up time window. White children tended to catch up with Black children in their pubertal status at the ages of between 13 and 15 years (**Figure 1B**). Notably, the magnitude of these effects estimated by PSM remained larger than that estimated by multiple regression, highlighting the sensitivity of PSM in detecting health differences over time.

We then investigated how such an obvious health difference between Black and White children could account for the other health differences observed during child development. For example, relative to White children, Black children have a higher BMI in the current dataset. This BMI difference between races can be partially explained by differences in pubertal status during child development. Controlling for pubertal status, either using PSM or multiple regression, can significantly diminish the difference in BMI between Black and White children by approximately 50% across 4-year longitudinal follow-ups (**Figure 1C & 1D**). These findings suggest that the magnitude of health differences between races may be inaccurately assessed if important health covariates such as pubertal status are not taken into account.

These findings further prompted us to investigate the extent to which pubertal status mediates health differences across diverse behavioral and neural measurements in adolescents (**Figure 2A**). We find that at baseline, pubertal status mediated 20 out of 40 (50%) behavioral, 192 out of 416 (46%) resting-state functional connectivity (rs-FC), and 173 out of 178 (97%) gray matter volume (GMV) measurements (**Figure 2B**). Among the 148 cortical GMVs, the strongest mediation effects emerge at the superior temporal gyrus and dorsal lateral as well as ventral medial prefrontal cortex (**Figure 2C**) – key brain regions previously implicated in social affective and cognitive development (Atzil et al., 2018; Fedorenko et al., 2024).

**Figure 2.**
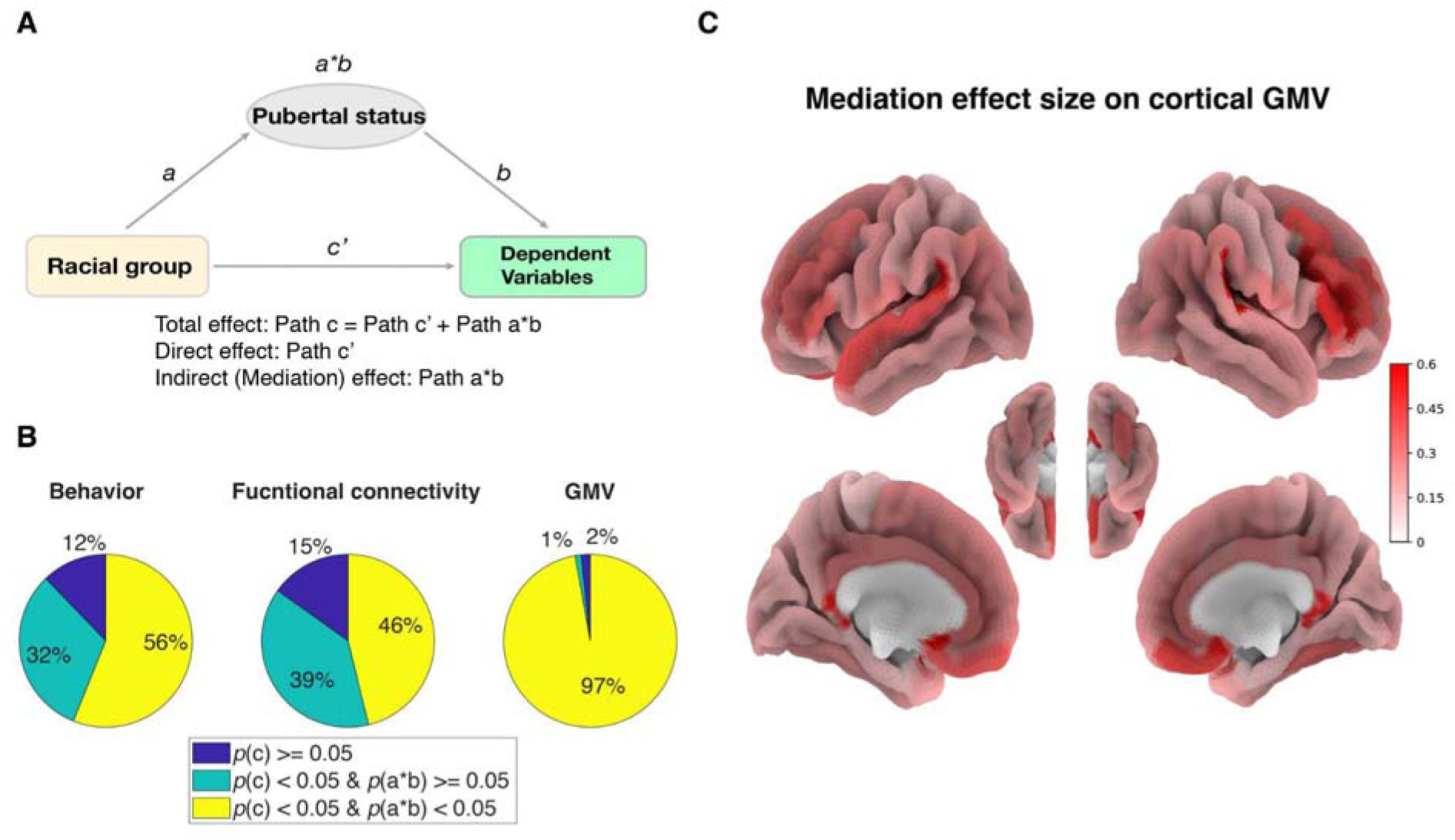
Mediation analyses. (**A**) The mediation diagram of pubertal status mediated the health differences between races. (**B**) Percentage of significant mediation effects (yellow) in 41 behavioral, 416 rs-FC, and 180 GMV measurements. (**C**) Illustration of mediation effect size on 148 cortical regions.

## Discussion

By leveraging a large dataset that captures the diverse characteristics of contemporary U.S. adolescents, our analyses suggest that PSM may offer a more sensitive covariate-control approach for assessing health-related differences between racial groups, as compared with traditional regression-based methods. Using this approach, we identified pubertal status as the most prominent difference between self-reported Black and White children. Controlling for pubertal status could diminish some of the health-related differences between these two racial groups. Overall, our study makes significant contributions to understanding health differences between racial groups.

First, methodologically, our analysis highlights the enhanced sensitivity of PSM in detecting health differences between Black and White children compared to conventional regression-based methods. The underperformance of multiple regression analyses can be attributed to multicollinearity between races and their associated covariates, such as SES. This shared variance among variables often violates the independence assumption of multiple regression, thereby diminishing the manifestation of health differences between races on certain dependent variables. In contrast, PSM enables the pairing of Black and White children based on key covariates, ensuring their equitable distribution between groups and minimizing the shared variance between groups and these key covariates. As a result, this approach enhances the sensitivity in uncovering and elucidating health differences between races.

Second, since the health differences between Black and White children are partially explained by pubertal status, controlling for this factor is helpful to obtain a more accurate understanding of health differences across races among U.S. adolescents. Puberty status is influenced by a combination of genetic, environmental, and socioeconomic factors. Earlier puberty is often linked to mental health issues, higher BMI, and experiences of discrimination (Argabright et al., 2022; Hoyt et al., 2020), particularly when children are unprepared for these changes due to a lack of timely puberty education (Hoyt et al., 2020). This educational gap is especially problematic for Black children, who tend to mature earlier than their White peers. Educators, clinicians, and parents play a crucial role in providing early puberty education, recognizing individual differences, and addressing potential biases related to physical development. It is important to note that controlling for puberty status has not been a common practice in previous studies investigating racial disparities/differences using the ABCD dataset (Dumornay et al., 2023; Giddens et al., 2022; Isaiah et al., 2022; Ryan et al., 2023). Additionally, our findings indicate that the differences in puberty status between racial groups gradually decrease as children grow older. Therefore, longitudinal studies must consider the potential effects of puberty status at different ages.

The current study has some limitations. First, as we only included self-reported Black and White children, our results may not be generalizable to other races/ethnicities. Nevertheless, the methods proposed in this study can be applied to address health inequalities among other racial or ethnic groups and assist in identifying key covariates that need to be considered. Second, as we used sex at birth as one of the basic covariates, we cannot compare sex differences in health-related outcomes. Consequently, the current study cannot detect whether there is an interaction effect between sex and race on pubertal status. Nonetheless, our current study unambiguously reveals systematic differences in health-related outcomes mediated by pubertal development between self-reported race groups. These effects could not be accounted for other covariates such as household income, parent education level, or other environmental factors, such as ADI.

In conclusion, these findings underscore the importance of using robust statistical methods like PSM to accurately capture health differences between self-reported races. It also suggests that researchers and policymakers need to consider factors like pubertal status when understanding and addressing health inequalities in adolescents.

## Key points

1. Understanding health differences between races in adolescents is important for addressing health inequalities
2. Conventional methods like multiple regression may not adequately control for covariates when health differences between races covary with common covariates
3. Propensity Score Matching offers better covariate control compared to traditional regression-based approaches
4. Puberty status emerged as a significant mediator of health differences between racial groups. This indicates that the timing of physical maturation plays a crucial role in health differences observed between self-reported Black and White children.
5. Researchers and policymakers need to consider factors like pubertal status when understanding and addressing health inequalities in adolescents.

## Data Availability

This work was based on a published dataset (Casey et al., 2018). The data analysis scripts have been deposited in https://github.com/nilsyang/Codes.

## Acknowledgments

This research was supported by the Intramural Research Program of the NIH, NINDS. We would like to thank Tina T. Liu for her constructive comments. We thank the ABCD consortium and NIH for providing the data for doing the research in this work. The ABCD Study is supported by the NIH and additional federal partners under award numbers U01DA041022, U01DA041028, U01DA041048, U01DA041089, U01DA041106, U01DA041117, U01DA041120, U01DA041134, U01DA041148, U01DA041156, U01DA041174, U24DA041123, and U24DA041147. A full list of supporters is available at https://abcdstudy.org/federal-partners. A listing of participating sites and a complete listing of the study investigators can be found at https://abcdstudy.org/principal-investigators. ABCD consortium investigators designed and implemented the study or provided data but did not necessarily participate in the analysis or writing of this report. This manuscript reflects the views of the authors and might not reflect the opinions or views of the NIH or ABCD consortium investigators. We are not paid to write this article by a pharmaceutical company or other agency.

## Author contributions

F.N.Y. designed research, analyzed data, and wrote the original paper; J.D. edited the paper and supervised the study; W.X. designed research, edited the paper, and supervised the study.

## Supplement

### Supplementary Figures

**Figure S1.**
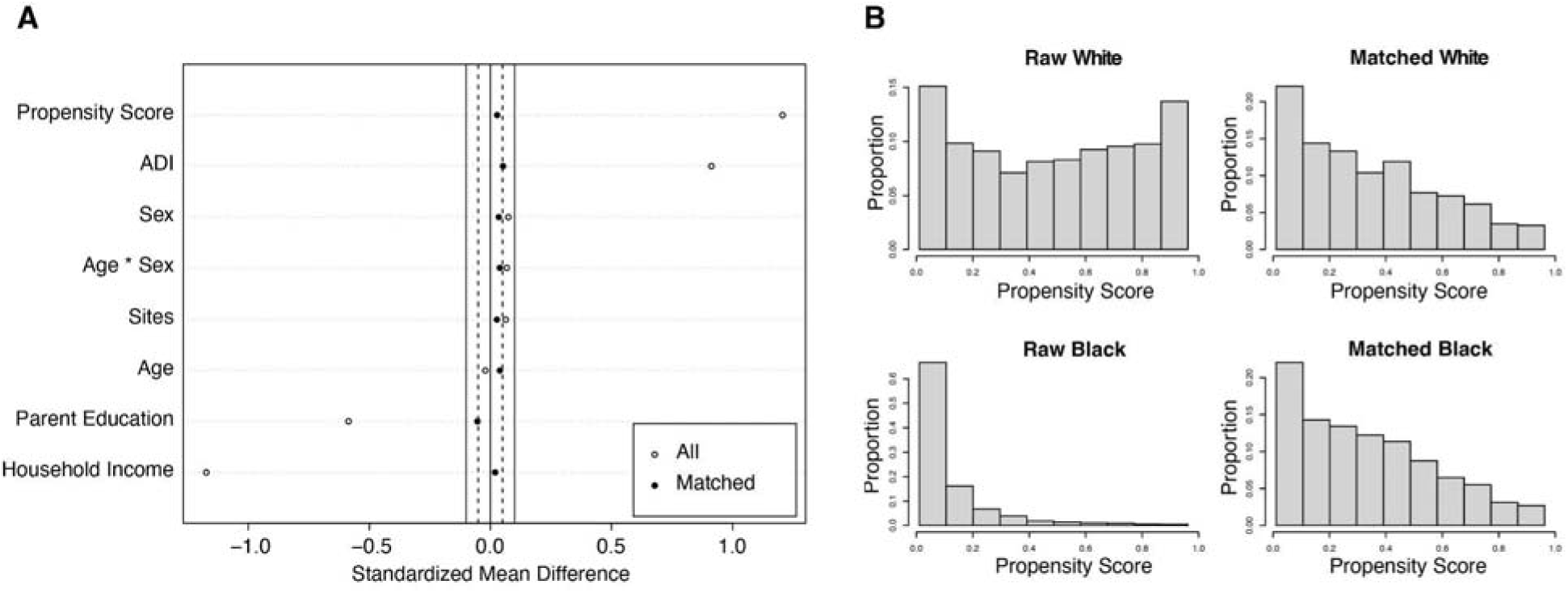
Propensity score matching controls for covariates. (A) The standardized mean differences of covariates between Black and White were reduced below 0.05. (B) The distributions of propensity scores after matching were similar between Black and White. Note: ADI = Area Deprivation Index.

